# SECONDARY STRESSORS AND THEIR PSYCHOSOCIAL IMPACTS ON HEALTHCARE STAFF: LESSONS FROM A QUALITATIVE SYSTEMATIC REVIEW FROM THE COVID-19 PANDEMIC IN THE UK

**DOI:** 10.1101/2024.04.16.24305910

**Authors:** Evangelos Ntontis, Richard Williams, Katarzyna Luzynska, Abigail Wright, Anastasia Rousaki

**Author notes:** Corresponding Author Dr Evangelos Ntontis, School of Psychology and Counselling, The Open University.

## Abstract

**Background:** Extreme events (e.g., floods and disease outbreaks) can overwhelm healthcare workers (HCWs) and healthcare systems. During the COVID-19 pandemic, high levels of distress and mental ill health were reported by HCWs.

**Aim:** To examine and synthesise research findings reported in the qualitative literature regarding the stressors, and their psychosocial impacts, that HCWs faced in the UK during the COVID-19 pandemic, and to provide lessons for future support.

**Method:** Qualitative articles were identified in EMBASE and OVID [pre-registered on PROSPERO: CRD42022304235]. Studies needed to have been published between January 2021 and January 2022 and to have examined the impact of COVID-19 on UK healthcare workers. We included 27 articles that represented the experiences of 2,640 healthcare workers, assessed their quality using the NICE criteria, and integrated their findings using thematic synthesis.

**Results:** Several secondary stressors were identified apart from the SARS-CoV-2 virus (the primary stressor), including lack of personal protective equipment, ineffective leadership and communication, high workloads, and problems stemming from uncertainty and a lack of knowledge. These stressors were related to various adverse psychosocial outcomes including worrying about oneself and others, fatigue, lack of confidence in oneself and in senior managers, impacts on teamwork, and feeling unappreciated or that one’s needs are not recognised.

**Conclusions:** Distress in HCWs proliferated due to the influence of secondary stressors. However, they can be modified to remove their negative effects. Consequently workforce planning should shift from focusing on individual change towards amending psychosocial environments in which healthcare staff work.

## INTRODUCTION

The COVID-19 pandemic, caused by the SARS-CoV-2 virus, led to large numbers of people being infected and high death rates across the globe. Healthcare workers were particularly affected because they were exposed to the virus through their work with infected people in addition to their exposure as members of the public. [1] Also, they witnessed other people being severely sick or dying and were working in extremely demanding environments. Thus, it is not surprising that multiple syntheses of evidence point to a high prevalence of reported symptoms of fatigue, distress, depression, anxiety, post-traumatic stress disorder, and reduced wellbeing. [1–6] Our aim in this qualitative systematic review is not to reiterate what previous researchers have shown regarding the psychosocial and mental health tolls that the COVID-19 pandemic took on healthcare workers in the UK as in other countries. Rather, we wish to move one step further and consider the antecedents, origins, and psychosocial impacts of the stressors that healthcare staff faced during the early stages of the pandemic in the UK and to reflect on lessons for future support during adverse challenges.

### Impacts of the COVID-19 Pandemic on Healthcare Staff

Existing research has highlighted the psychosocial impacts of the COVID-19 pandemic on healthcare staff across the world. Two meta-analyses of the impact of the pandemic on HCWs showed that a large percentage reported experiencing symptoms of depression, anxiety, and insomnia.[3,5] Similar systematic meta-analytic examinations of the mental health problems experienced by HCWs showed that the latter reported experiencing symptoms of post-traumatic stress disorder, anxiety, depression, insomnia, burnout, and distress.[2,4] These effects can be exacerbated by particular risk factors such as being female, working in the frontlines, being a nurse, being younger, having less social support, or experiencing more occupational stressors (e.g., significant workloads, lacking protective equipment, working in shared spaces) among others.[1,3,5,7] These results are far from surprising. In general populations that experience extreme events, people with higher levels of exposure, women, ethnic minorities, less affluent, and people with fewer psychosocial resources or people with previous experiences of mental health problems are more likely to demonstrate adverse psychological outcomes.[8]

Crucially, Norris and colleagues [8] list secondary stressors as one of the risk factors associated with the prevalence of psychological problems. In contrast to *primary stressors* that are inherent in extreme events such as viral infections, floodwaters, or fires,[9] secondary stressors have been defined as *‘1. Social factors and people’s life circumstances (that include the policies, practices, and social, organisational, and financial arrangements) that exist prior to and impact them during the major incident; and/or 2. Societal and organisational responses to an incident or emergency’.* [10,11] Examples of secondary stressors in extreme events such as floods, hurricanes, and earthquakes include problems in claiming insurance payments, insufficient housing, miscommunication, poor living and working conditions that persist, and people’s disconnection from healthcare and other services on which they rely.[12–15]

### The Workforce Context in the UK

Considering that secondary stressors primarily reflect organisational deficiencies and ineffective responses to extreme events that can harm workers, we briefly describe the situation affecting the healthcare workforce in the UK and how it can promote distress. Beneficial scientific advances in healthcare have contributed to a chronic imbalance between supply and demand in a setting of severe budgetary limitations. This is, if anything, worsening.[16] The NHS workforce has faced chronic strain over many years, with workload pressures continuing to grow, and the imbalances becoming progressively harder to meet in the last decade particularly.[17]

Before the pandemic, retention, recruitment, and mental health challenges were exacerbating long-term problems with working conditions.[18] Yet, it is also evident that some of the causes of pressure were not only fiscal but resulted from stressors that were not adequately recognised or dealt with. In 2018, a report by the UK’s General Medical Council presented evidence regarding the sources of pressure faced by doctors, pointing towards issues related to workload, staffing, work-life balance, and lack of support among others.[17] The same report stated that the NHS was ‘at a critical juncture’ (p. 24). The following year, a report by the British Medical Association [19] pointed to similar stressors faced by healthcare staff including poor work-life balance, understaffing, lack of time for professional development, inability to provide care perceived as adequate, problematic hierarchies and bullying, or lacking basic amenities and adequate breaks among others.

The aforementioned problems persisted during the COVID-19 pandemic, and, particularly during its earlier stages, intensified the impact on HCWs’ wellbeing, stress, fatigue, and burnout. More recently, Oeppen et al. have called for the NHS to do more to prevent fatigue in healthcare staff because the needs of staff have not ended or stopped rising as the additional pressures from COVID-19 have reduced.[20] This suggests that a closer look into the psychosocial impacts of particular organisational arrangements and responses in the NHS is crucial if HCWs are to be supported adequately in both routine care and future extreme events.

Reflecting on lessons for the future is important for at least three reasons. First, it was not the COVID-19 pandemic that first created unsustainable demands in the UK’s healthcare system; the NHS had been under huge strain for a long time before the pandemic emerged and various of the stressors that HCWs faced were already prevalent.[21] Second, many of the stressors that healthcare staff experienced resulted from sub-optimal responses to the pandemic by governments and/or healthcare systems at the point when the pandemic emerged and during subsequent waves. Third, considering that the climate crisis is increasing the frequency and intensity of extreme events,[22, 23] as well as recent dismantling of public healthcare services in neoliberal economies,[24,25] healthcare systems and the staff working in them are very likely to face more demands and strain in the future. What all three points share is that the stressors described are not only tractable due to their being rooted in particular systems and practices, but they are also amenable to change through both pre-disaster preparedness activities and adequate institutional responses when incidents occur.

When attempting to address HCWs’ wellbeing, psychosocial, and mental healthcare needs,[18] it becomes clear that solely focusing on enhancing people’s personal resilience remains far from adequate. In the face of severe and long-lasting impacts of systemic and institutional deficiencies, it has became paramount that what is necessary continues to be a stronger focus on how particular institutional structures exacerbate and sustain distress but also, more positively, on how social support and the physical, psychosocial and moral environments within which healthcare workers have found themselves can buffer negative experiences.[26–28]

### The Study Reported in This Paper

This paper reports a systematic review developed from a broader project commissioned by NHS England that concerned scoping the literature published in the UK between 2021 and early 2022 regarding the impacts of COVID-19 on healthcare workers during the earlier stages of the pandemic. The consistently high quality of the qualitative papers and the overarching theoretical significance of their findings led us to develop and conduct the pre-registered qualitative systematic review that we present here.

## METHOD

### Search Strategy and Selection Criteria

The design and reporting of our review were informed by the Preferred Reporting Items for Systematic Reviews and Meta-Analysis (PRISMA) statement (http://www.prisma-statement.org/). Our review was registered with and met the criteria set by PROSPERO, the international prospective register of systematic reviews (CRD42022304235; registration accepted on 31 January 2022). PROSPERO accepts registrations of systematic reviews before data screening or extraction begins. Consequently, the databases were test-searched on 18 January 2022 and then on 29 January 2022.

AW searched OVID and EMBASE, two major scientific databases in the social, psychological, and health sciences. Our search terms included variations of certain keywords such as: COVID-19; wellbeing; distress; psychological; psychosocial; mental health; staff; doctor; allied health; nurse; NHS; social care; consultant; medical staff; United Kingdom. We only included articles published in English that referred to the United Kingdom and were published between January 2021 and January 2022. Non-empirical reports, grey literature, blogs, or opinions pieces were excluded. The keywords used as search terms in the databases were tailored appropriately for each database.

### Selection Procedure, Screening, Data Extraction and Quality Assessment

The literature search returned 2,277 studies, the titles, abstracts, and other supporting information of which were entered in a spreadsheet. Of these, 437 were duplicate records and 188 studies were published outside the pre-specified year range (there was some overlap between those two categories) and in total we removed 608 records before the screening stage. Subsequently, two authors (KL,EN) screened the titles and abstracts of the remaining 1,669 articles, excluding those not relevant to our aims (e.g., papers reporting quantitative studies). This led to 1,628 papers being removed. In cases of doubt, the procedure was for the reviewer to discuss with the other reviewer and, if disagreement occurred, a third author (RW) was to be consulted. The latter course was not required. The screening process started on 2 February 2022 and we extracted the data on 8 March, 2022. Following the screening stage, we proceeded to data retrieval. At this stage, we sought to retrieve both qualitative as well as mixed-methods papers, since we found that the latter include a qualitative aspect. We sought to retrieve 41 papers but could not retrieve 4; 1 was qualitative, and 3 used mixed methods. Our overall search and selection process is shown in Figure 1.

**Figure 1.**
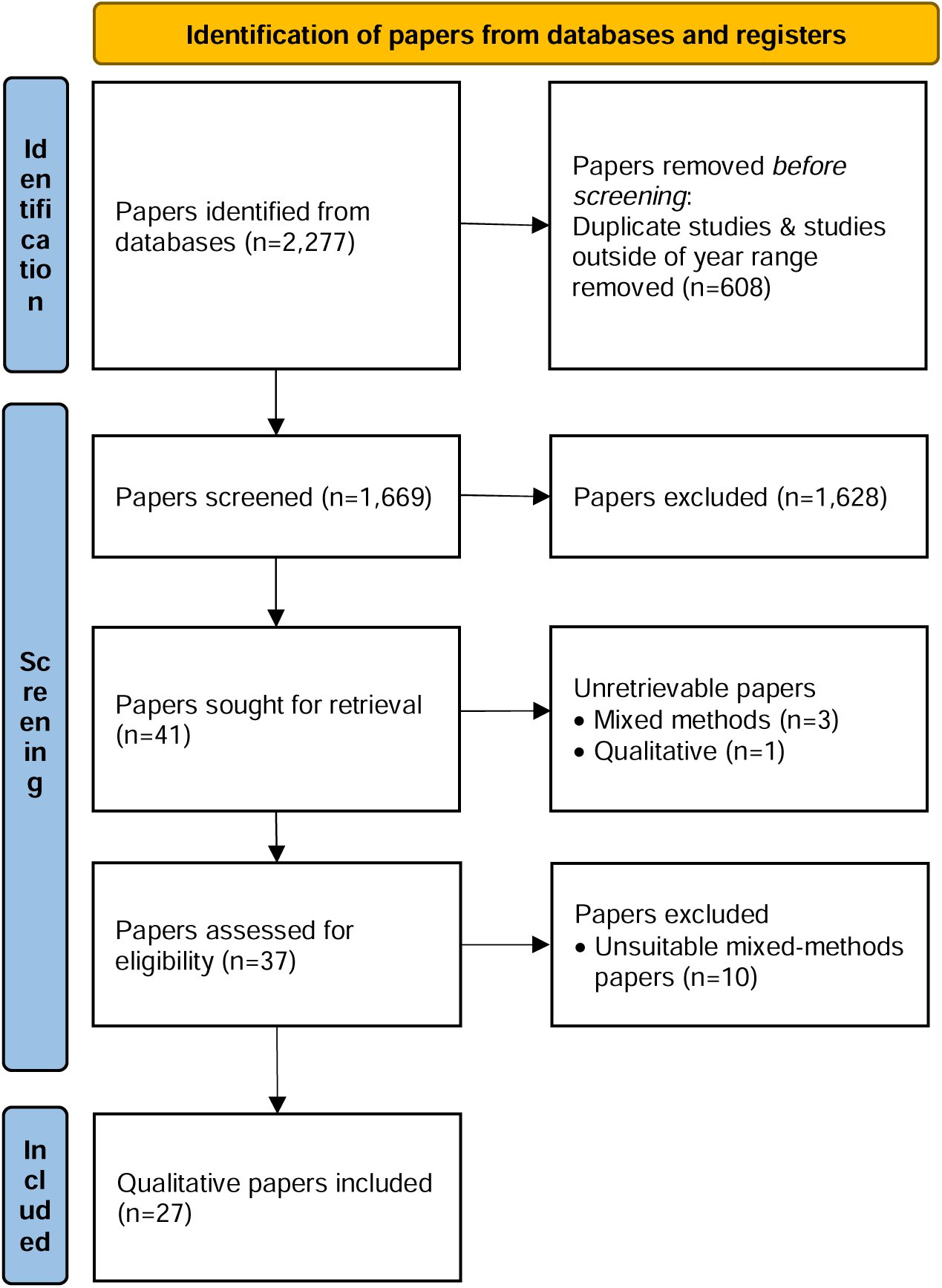
PRISMA flow diagram of search and selection process.

Upon re-inspecting the literature included in this study in mid-2024, we were able to identify 1 previously unretrievable qualitative study (a pre-print that was eventually published around 7 months after our database search took place), and 1 mixed-methods study. However we decided not to include these papers because they had not been published when our search was taking place and and, therefore lie outside the terms of the protocol.

We assessed the 37 papers that we retrieved. We decided to not include 10 papers reporting mixed-methods studies because those papers did not report in-depth analyses of the qualitative data and did not satisfy the quality criteria reported below. Many reported the results of quantitative questionnaires, which included open-ended questions at the end and, in some cases, presented isolated quotes, or descriptive summaries of these quotes. However, they did not reach the depth required for stand-alone qualitative analyses. Nevertheless, we did examine separately the qualitative aspects of these mixed-methods papers and found that their findings were in line with the results we report in this paper.

For transparency, the 10 mixed-methods papers are listed in the Supplementary Materials. We have no reason to believe that not including them has altered our findings.

We used the quality appraisal checklist endorsed by the National Institute for Health and Care Excellence (NICE)[29] to assess the quality of the 27 qualitative papers that comprised the final dataset of this systematic review. NICE has a range of criteria against which it assesses research studies, such as the appropriateness of a qualitative design, clarity of study aims and data collection processes, thorough descriptions of source population, rigour of the analysis and richness of the data presented, reliability of the analysis, appropriateness of conclusions and their grounding on the data, ethical issues, quality control, and reflexivity among others. All papers were assessed by three team members (KL,EN,AR) and were given a score based on the these criteria. There were no disagreements between the team members. Most papers were judged as being of very high quality as they fullfulled the suggested criteria. We identified some minor limitations in 4 papers (papers m, t, y, aa in Table 1; e.g., the discussion section in one paper did not adequately reflect the results; some papers did not include reflexivity statements). When considering whether to disregard these papers, we took into account other strengths (e.g., well-written analyses, clear aims and sampling procedures, quotes used to illustrate their points) as well as the findings being in line with other studies; thus we decided to keep them in the final synthesis. A list of papers, their aims and characteristics, and quality control measures is in Table 1.

**Table 1:**
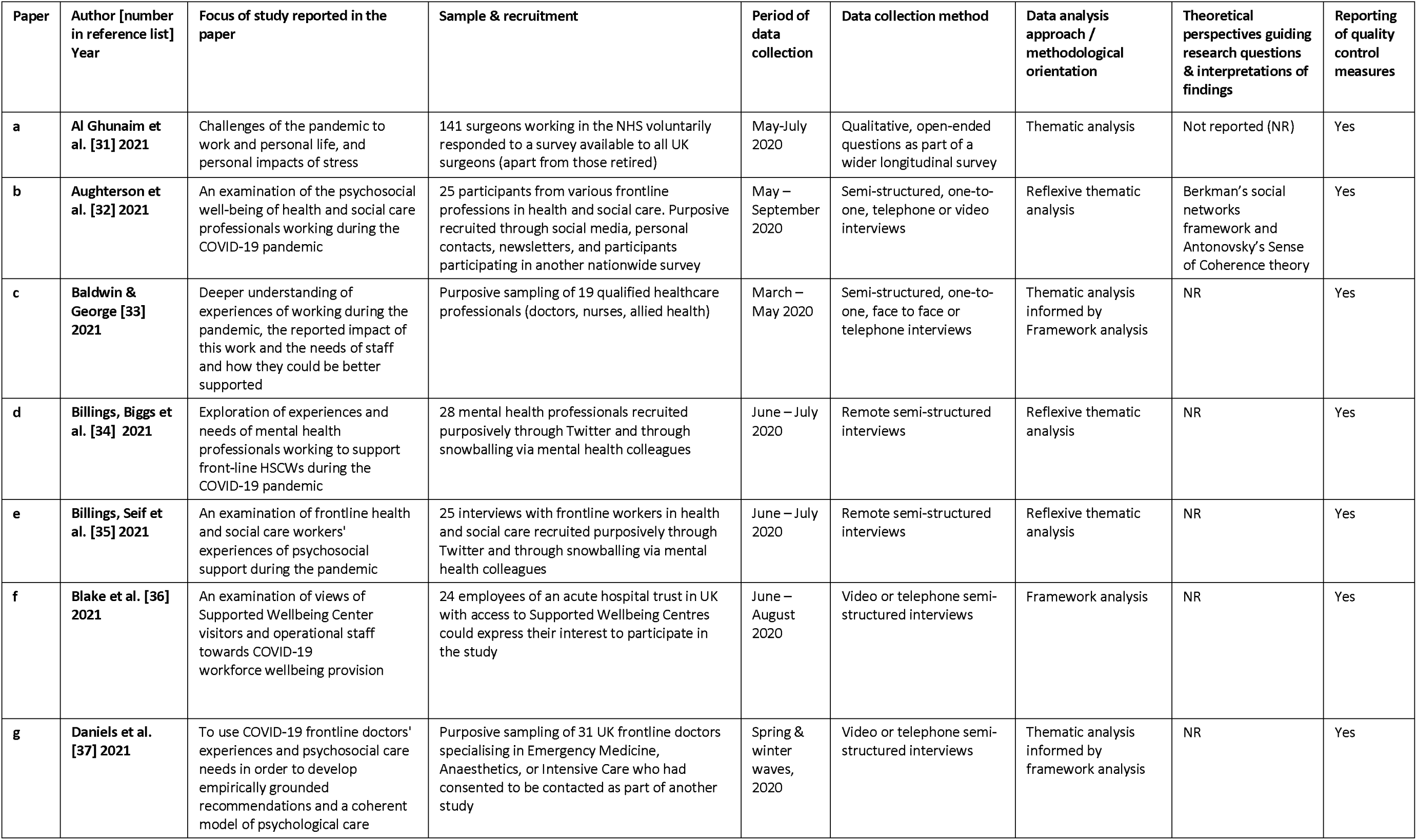
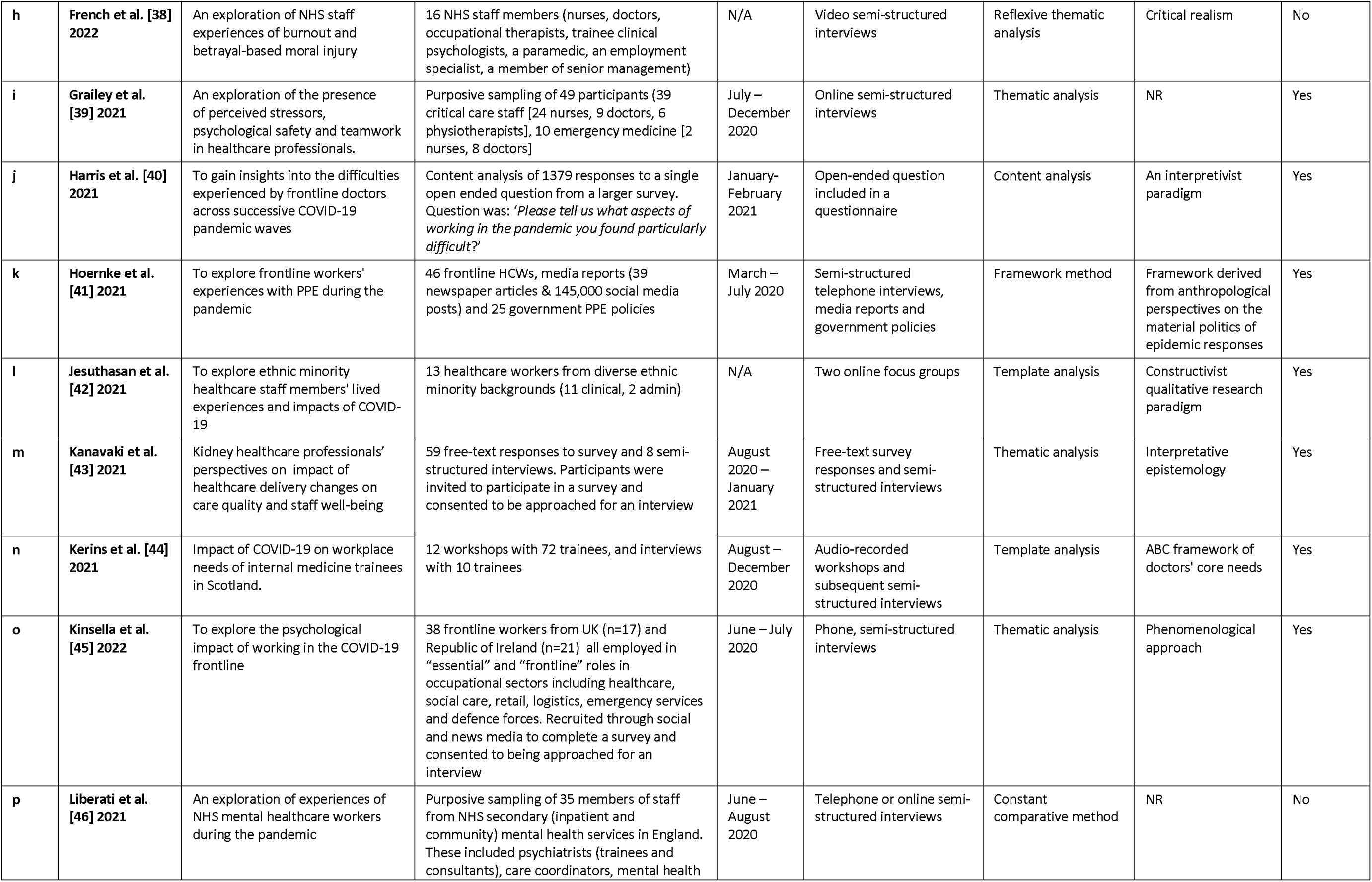
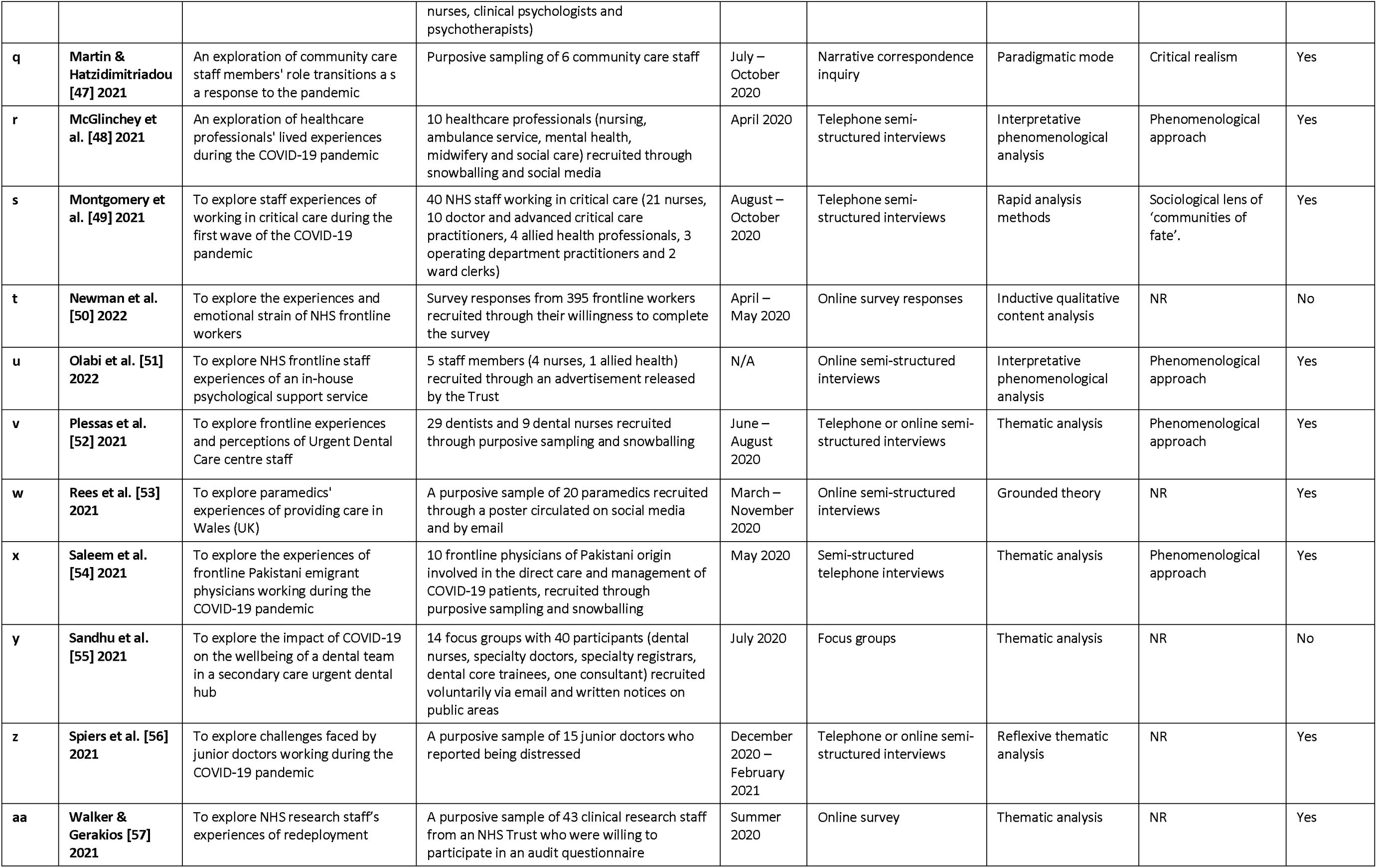
The papers reporting qualitative studies that were reviewed systematically.

| Paper | Author [number in reference list] Year | Focus of study reported in the paper | Sample & recruitment | Period of data collection | Data collection method | Data analysis approach / methodological orientation | Theoretical perspectives guiding research questions & interpretations of findings | Reporting of quality control measures |
| --- | --- | --- | --- | --- | --- | --- | --- | --- |
| a | Al Ghunaim et al. [31] 2021 | Challenges of the pandemic to work and personal life, and personal impacts of stress | 141 surgeons working in the NHS voluntarily responded to a survey available to all UK surgeons (apart from those retired) | May-July 2020 | Qualitative, open-ended questions as part of a wider longitudinal survey | Thematic analysis | Not reported (NR) | Yes |
| b | Aughterson et al. [32] 2021 | An examination of the psychosocial well-being of health and social care professionals working during the COVID-19 pandemic | 25 participants from various frontline professions in health and social care. Purposive recruited through social media, personal contacts, newsletters, and participants participating in another nationwide survey | May – September 2020 | Semi-structured, one-to-one, telephone or video interviews | Reflexive thematic analysis | Berkman's social networks framework and Antonovsky's Sense of Coherence theory | Yes |
| c | Baldwin & George [33] 2021 | Deeper understanding of experiences of working during the pandemic, the reported impact of this work and the needs of staff and how they could be better supported | Purposive sampling of 19 qualified healthcare professionals (doctors, nurses, allied health) | March – May 2020 | Semi-structured, one-to-one, face to face or telephone interviews | Thematic analysis informed by Framework analysis | NR | Yes |
| d | Billings, Biggs et al. [34] 2021 | Exploration of experiences and needs of mental health professionals working to support front-line HSCWs during the COVID-19 pandemic | 28 mental health professionals recruited purposively through Twitter and through snowballing via mental health colleagues | June – July 2020 | Remote semi-structured interviews | Reflexive thematic analysis | NR | Yes |
| e | Billings, Seif et al. [35] 2021 | An examination of frontline health and social care workers' experiences of psychosocial support during the pandemic | 25 interviews with frontline workers in health and social care recruited purposively through Twitter and through snowballing via mental health colleagues | June – July 2020 | Remote semi-structured interviews | Reflexive thematic analysis | NR | Yes |
| f | Blake et al. [36] 2021 | An examination of views of Supported Wellbeing Center visitors and operational staff towards COVID-19 workforce wellbeing provision | 24 employees of an acute hospital trust in UK with access to Supported Wellbeing Centres could express their interest to participate in the study | June – August 2020 | Video or telephone semi-structured interviews | Framework analysis | NR | Yes |
| g | Daniels et al. [37] 2021 | To use COVID-19 frontline doctors' experiences and psychosocial care needs in order to develop empirically grounded recommendations and a coherent model of psychological care | Purposive sampling of 31 UK frontline doctors specialising in Emergency Medicine, Anaesthetics, or Intensive Care who had consented to be contacted as part of another study | Spring & winter waves, 2020 | Video or telephone semi-structured interviews | Thematic analysis informed by framework analysis | NR | Yes |
| h | French et al. [38] 2022 | An exploration of NHS staff experiences of burnout and betrayal-based moral injury | 16 NHS staff members (nurses, doctors, occupational therapists, trainee clinical psychologists, a paramedic, an employment specialist, a member of senior management) | N/A | Video semi-structured interviews | Reflexive thematic analysis | Critical realism | No |
| i | Grailey et al. [39] 2021 | An exploration of the presence of perceived stressors, psychological safety and teamwork in healthcare professionals. | Purposive sampling of 49 participants (39 critical care staff [24 nurses, 9 doctors, 6 physiotherapists], 10 emergency medicine [2 nurses, 8 doctors]) | July – December 2020 | Online semi-structured interviews | Thematic analysis | NR | Yes |
| j | Harris et al. [40] 2021 | To gain insights into the difficulties experienced by frontline doctors across successive COVID-19 pandemic waves | Content analysis of 1379 responses to a single open ended question from a larger survey. Question was: 'Please tell us what aspects of working in the pandemic you found particularly difficult?' | January-February 2021 | Open-ended question included in a questionnaire | Content analysis | An interpretivist paradigm | Yes |
| k | Hoernke et al. [41] 2021 | To explore frontline workers' experiences with PPE during the pandemic | 46 frontline HCWs, media reports (39 newspaper articles & 145,000 social media posts) and 25 government PPE policies | March – July 2020 | Semi-structured telephone interviews, media reports and government policies | Framework method | Framework derived from anthropological perspectives on the material politics of epidemic responses | Yes |
| l | Jesuthasan et al. [42] 2021 | To explore ethnic minority healthcare staff members' lived experiences and impacts of COVID-19 | 13 healthcare workers from diverse ethnic minority backgrounds (11 clinical, 2 admin) | N/A | Two online focus groups | Template analysis | Constructivist qualitative research paradigm | Yes |
| m | Kanavaki et al. [43] 2021 | Kidney healthcare professionals' perspectives on impact of healthcare delivery changes on care quality and staff well-being | 59 free-text responses to survey and 8 semi-structured interviews. Participants were invited to participate in a survey and consented to be approached for an interview | August 2020 – January 2021 | Free-text survey responses and semi-structured interviews | Thematic analysis | Interpretative epistemology | Yes |
| n | Kerins et al. [44] 2021 | Impact of COVID-19 on workplace needs of internal medicine trainees in Scotland. | 12 workshops with 72 trainees, and interviews with 10 trainees | August – December 2020 | Audio-recorded workshops and subsequent semi-structured interviews | Template analysis | ABC framework of doctors' core needs | Yes |
| o | Kinsella et al. [45] 2022 | To explore the psychological impact of working in the COVID-19 frontline | 38 frontline workers from UK (n=17) and Republic of Ireland (n=21) all employed in "essential" and "frontline" roles in occupational sectors including healthcare, social care, retail, logistics, emergency services and defence forces. Recruited through social and news media to complete a survey and consented to being approached for an interview | June – July 2020 | Phone, semi-structured interviews | Thematic analysis | Phenomenological approach | Yes |
| p | Liberati et al. [46] 2021 | An exploration of experiences of NHS mental healthcare workers during the pandemic | Purposive sampling of 35 members of staff from NHS secondary (inpatient and community) mental health services in England. These included psychiatrists (trainees and consultants), care coordinators, mental health nurses, clinical psychologists and psychotherapists) | June – August 2020 | Telephone or online semi-structured interviews | Constant comparative method | NR | No |
| q | **Martin & Hatzidimitriadou [47] 2021** | An exploration of community care staff members' role transitions as a response to the pandemic | Purposive sampling of 6 community care staff | July – October 2020 | Narrative correspondence inquiry | Paradigmatic mode | Critical realism | Yes |
| r | **McGlinchey et al. [48] 2021** | An exploration of healthcare professionals' lived experiences during the COVID-19 pandemic | 10 healthcare professionals (nursing, ambulance service, mental health, midwifery and social care) recruited through snowballing and social media | April 2020 | Telephone semi-structured interviews | Interpretative phenomenological analysis | Phenomenological approach | Yes |
| s | **Montgomery et al. [49] 2021** | To explore staff experiences of working in critical care during the first wave of the COVID-19 pandemic | 40 NHS staff working in critical care (21 nurses, 10 doctor and advanced critical care practitioners, 4 allied health professionals, 3 operating department practitioners and 2 ward clerks) | August – October 2020 | Telephone semi-structured interviews | Rapid analysis methods | Sociological lens of 'communities of fate'. | Yes |
| t | **Newman et al. [50] 2022** | To explore the experiences and emotional strain of NHS frontline workers | Survey responses from 395 frontline workers recruited through their willingness to complete the survey | April – May 2020 | Online survey responses | Inductive qualitative content analysis | NR | No |
| u | **Olabi et al. [51] 2022** | To explore NHS frontline staff experiences of an in-house psychological support service | 5 staff members (4 nurses, 1 allied health) recruited through an advertisement released by the Trust | N/A | Online semi-structured interviews | Interpretative phenomenological analysis | Phenomenological approach | Yes |
| v | **Plessas et al. [52] 2021** | To explore frontline experiences and perceptions of Urgent Dental Care centre staff | 29 dentists and 9 dental nurses recruited through purposive sampling and snowballing | June – August 2020 | Telephone or online semi-structured interviews | Thematic analysis | Phenomenological approach | Yes |
| w | **Rees et al. [53] 2021** | To explore paramedics' experiences of providing care in Wales (UK) | A purposive sample of 20 paramedics recruited through a poster circulated on social media and by email | March – November 2020 | Online semi-structured interviews | Grounded theory | NR | Yes |
| x | **Saleem et al. [54] 2021** | To explore the experiences of frontline Pakistani emigrant physicians working during the COVID-19 pandemic | 10 frontline physicians of Pakistani origin involved in the direct care and management of COVID-19 patients, recruited through purposive sampling and snowballing | May 2020 | Semi-structured telephone interviews | Thematic analysis | Phenomenological approach | Yes |
| y | **Sandhu et al. [55] 2021** | To explore the impact of COVID-19 on the wellbeing of a dental team in a secondary care urgent dental hub | 14 focus groups with 40 participants (dental nurses, specialty doctors, specialty registrars, dental core trainees, one consultant) recruited voluntarily via email and written notices on public areas | July 2020 | Focus groups | Thematic analysis | NR | No |
| z | **Spiers et al. [56] 2021** | To explore challenges faced by junior doctors working during the COVID-19 pandemic | A purposive sample of 15 junior doctors who reported being distressed | December 2020 – February 2021 | Telephone or online semi-structured interviews | Reflexive thematic analysis | NR | Yes |
| aa | **Walker & Gerakios [57] 2021** | To explore NHS research staff's experiences of redeployment | A purposive sample of 43 clinical research staff from an NHS Trust who were willing to participate in an audit questionnaire | Summer 2020 | Online survey | Thematic analysis | NR | Yes |

The papers were read by three members of the team (EN,KL,AR). KL and AR manually extracted information on the papers’ authors, year of publication, period of data collection, study aims, methodology and/or theoretical frameworks, sample, and reporting of quality control. These data were imported into an Excel file and became the basis for our analysis. EN re-read the papers independently and cross-checked the analyses against the data extracted by KL and AR to ensure consistency and accuracy.

The authors of the 27 papers used a range of data collection approaches such as interviews, focus groups, and online surveys with open-ended questions. They also used a range of analytical approaches (e.g., thematic analysis, content analysis, interpretative phenomenological analysis [IPA]) and theoretical frameworks). The samples spanned the experiences of a range of health and social care disciplines including doctors, nurses, psychologists, dentists, and occupational therapists.

### Thematic Synthesis

We followed the approach of Thomas and Harden when conducting the thematic synthesis by, first, coding the findings of the primary studies, then creating descriptive themes, and finally establishing analytical themes. [30]

The first stage involved going through the dataset and labeling evidence of the themes and their summaries using core keywords that, on the one hand, encapsulated their meaning and, on the other hand, allowed us to find commonalities across the dataset (e.g., personal protective equipment [PPE], or leadership were codes used to label elements identified in the data and to differentiate them from one another). Subsequently, descriptive themes were created which identified commonalities across the various codes (e.g., fear of contracting COVID-19 or lack of PPE) that eventually incorporated the psychosocial impacts of stressors and led to our final analytical themes. All authors had input into the analysis and writing the paper was led by EN and RW.

## RESULTS

The papers that we reviewed in detail are listed in Table 1. In the text, we refer to them by the letter that appears in the left-hand column.

Our analysis generated five themes; all reflect processes that occurred during the earlier stages of the pandemic. The first theme appraises concerns and negative emotions and experiences stemming directly from the the illness -COVID-19 - caused by the virus SARS-CoV-2.. The second theme describes challenges related to PPE, and the third theme addresses leadership and communication problems. The fourth theme reflects uncertainty and lack of knowledge and how they were outcomes of organisational features of staff members’ institutions, and the fifth theme focuses on workload problems and their psychosocial impacts.

### Theme 1: Fear and Worry about The Direct Impact of COVID-19

As expected, in many instances, participants expressed their fear of contracting COVID-19 due to their being in an environment that exposed them to patients who tested positive for the virus.[c,f,h,i,l,n,o,r,s,t,v,w,y,z] Grailey et al.[i] illustrate this fear through an account from a senior staff nurse who stated that:

> *It was very much felt that the people who were going in could potentially be in harm’s way.*

A similar exemplary account from a trainee is provided by Kerins et al.[n]:

> *Worried about spreading it … or worried about catching it … or worried about spreading it to their family or bringing it into the hospital.*

The latter quote provides insights into the multiple layers of concerns that participants experienced and reported at the onset of the pandemic. For example, due to the nature of their jobs, participants were afraid not only of contracting COVID-19 themselves, but also of transmitting it to family members and friends, thus raising concerns about the safety of people other than themselves,[a,b,e,f,l,s,v,w,y] some of whom could have underlying health problems.[b,s,w] Al-Ghunaim et al.[a] provide one such account from a surgeon:

> *Fear of bringing the virus home and infecting my family and my mother-in-law with lung cancer.*

The authors of some papers raised concerns regarding the psychosocial toll of HCWs witnessing so many patients being critically ill or dying,[f,o,r,s,z] and witnessing frontline workers dying.[f,s,w] Participants also reported fear of the unknown and a sense of uncertainty stemming from the pandemic during its earlier stages,[b,I,n,t] and feelings such as anxiety,[c,m,s,w,y] isolation,[e,i,o,y] despair and grief.[f,t,z] Kanavaki et al.[m] depict the psychosocial toll of the pandemic through a doctor’s account which illustrates not only the psychological but also the social impacts of COVID-19 on the lives of health and social care workers:

> *There is anxiety relating to uncertainty and a demoralisation as so many planned activities are cancelled and contact with friends and family is reduced.*

The various elements summarised thus far provide a picture of the ways in which awareness of SARS-CoV-2 brought distress to healthcare workers during the onset of COVID-19 through a range of negative experiences, feelings and psychosocial impacts. Despite COVID-19 directly causing distress for participants, as is evident from the data and the wider literature, the effects of the virus were exacerbated by their interaction with a range of other stressors rooted in pre-pandemic systemic issues as well as those introduced by certain ineffective responses to the pandemic. We present this matter in the themes that follow.

### Theme 2: Problems Related to Personal Protective Equipment and their Negative Psychosocial Outcomes

One central factor that was commonly reported as contributing to staff members’ experiences of distress was lack of access to personal protective equipment (PPE) that raised difficulties in delivering care.[a,c,f,i,k,r,t,v,x] As Baldwin and George [c] quote:

> *… police officers don’t go out without a stab vest; firemen don’t go out without wearing the full protective gear … why are healthcare staff any different? Why are we not provided with the appropriate [PPE].*

During the onset of the pandemic, there was widespread dissatisfaction regarding the lack of PPE. However, the lack of PPE was not a problem directly attributable to the pandemic. Rather, it was both a pre-existing deficiency inherent in limitations in preparation and planning in the healthcare system in England as well as problematic organisational responses to the emergent extreme event. In some cases, participants reported that PPE was available, but they were worried about both its quality and quantity.[j,k,r,w] These concerns were reported as distressing. One reason was cited by participants who acknowledged the limited nature of these resources, and did not take breaks because, by doing so, they would have to discard parts of their equipment, which was even more necessary early in the pandemic. Concerns about PPE led to increased fatigue. We present an extract from a longer quote as it appears in McGlinchey et al.[r]:

> *I basically have a drink of water and then put on the equipment again, because other people are waiting for their breaks, and go back in … Once you are in, you can’t really come out just to go to the toilet because it wastes PPE … you are starting to get dehydrated because you can’t drink …*

Participants also reported a lack of adoption of fit testing at the early stages of the pandemic,[v,y] and feelings of fear that stemmed from an associated perceived lack of safety due to the absence of fit testing.[c,w] According to a quote presented by Rees et al.[w]:

> *… really worried because I knew the masks didn’t fit me properly, so I was anxious and I felt a bit demotivated to be in work, that I didn’t want to be there because every day I was going in and it was a permanent risk really.*

Another related issue was the lack of training in how to properly use PPE, even when the latter was available, which made staff feel less confident in carrying out their duties in a manner that was safe for them and safe for patients and colleagues.[k,y] According to a quote presented by Hoernke et al.[k]:

> *haven’t had any training … some other nurses have been trained to use ventilators but there hasn’t been any PPE training or anything else at all.*

Overall, many staff did not feel protected against the pandemic during its early stages,[a,f] and were afraid of transmitting it to their families and friends,[f] revealing one of many potential connections between a stressor rooted in institutional and organisational problems (in this case the lack of PPE) and its interaction with the direct psychosocial effects of the extreme event (the fear of contracting the virus) that was often accentuated for members of staff from black and minority ethnic cultures.[f]

PPE was reported as causing personal discomfort and this became a factor that affected staff-patient relationships and staff team dynamics. Lengthy working in PPE caused discomfort and fatigue,[b,i,k,w,y] and changing from used to new equipment frequently took time, and became a burden.[a] Moreover, necessary use of PPE became a barrier to staff becoming familiar and communicating with other team members,[i,k,s,v] made it harder for them to communicate with patients,[b] and created dilemmas between effective risk infection and human contact,[p] by limiting: visual and auditory cues; the ability to recognise and communicate with others; and coordinating and executing various tasks.[a,i,k,s] Overall, it was reported as having a negative impact on teamwork.[i] As Al-Ghunaim et al [a] quote:

> *PPE makes it difficult for patients to hear you and see your non-verbal response.*

PPE-related matters were the causes of wider problems in workplaces. Recurrently changing guidance and seemingly never-ending changes to levels of PPE required for different healthcare processes did not inspire workers’ confidence in safety and senior managers.[w,y]. According to a quote from Rees et al.[w]:

> *you have been thrown information constantly, there’s updates after updates after updates, things are changing near enough I wouldn’t say hourly but frequently changing. Yes, you are probably being suitably informed, but it is overwhelming.*

Lack of organisational trust and leadership problems were common and we consider these topics next.

### Theme 3: Problems With Leadership and Communications

Problems in leadership and in communications were two interconnected matters that were reported frequently by participants as causing significant distress among HCWs. For instance, many participants mentioned that they were unclear about rapidly changing guidelines leading to confusion.[c,j,k,o,t,v,w,y,aa] Plessas et al.[v], for example, present a quote:

> *Nationally, I think things could have been done better, especially from the CDO’s [Chief Dental Officer] office; there were a lot of confusing messages coming out …*

A similar issue was staff receiving too much information from too many different sources without proper control over what was being disseminated, leading to staff members’ inability to trust both the sources as well as the information disseminated or to conflicting information reaching the clinical services.[e,s,t,w,aa]

On other occasions, a common pattern in the papers was that of participants reporting that there was a lack of communication from staff in more senior jobs,[c,g,j,p,r,v,aa] coupled with information often not being disseminated to teams.[c,aa] Walker and Gerakios [aa] present a quote:

> *Difficult to keep up with changes if they are not communicated effectively.*

Overall, these patterns of findings point us to the distressing experiences that staff faced due to poor communication with senior managers, particularly in the first and second waves of the pandemic, that became an obstacle to the former being able to carry out their jobs in an effective and safe manner.

Leadership problems were more broadly implicated in staff members’ negative psychosocial experiences and especially if they were superimposed on other problems. For instance, participants reported receiving no support or feeling unsupported due to having no managers on the ground, with more senior people only contacting them through email.[c,f,h,j,l,m,t] Baldwin and George[c] quote a staff member saying:

> *I have never seen any of the management people in the PPE to come in and to see what happens.*

Problems experienced as stemming from managers led to participants feeling unappreciated, undervalued, and misunderstood.[g,n,r,t]A quote from McGlinchey et al.[r] illustrates this matter:

> *We work in a caring profession, but you don’t feel cared for.*

A sense of unfairness emerged from some participants because of their having to carry out significantly more tasks and of having to cope with massive workloads while receiving the same salary.[p,v] Disbanding rest areas and removing free parking and meals (all set up by some employers very early on) also led to some participants feeling undervalued.[e,n,p] Many participants felt ignored or forgotten by managers.[b,f,g] They experienced a sense of abandonment and that they were seen as disposable, throwaway resources and purely as numbers despite managers being aware of the dangers that staff were facing.[c,h] French et al.[h] illustrate this with a powerful quote from a nurse:

> *If I die, they don’t care. It doesn’t matter if [they] get like, you know, 600 nurses have died from COVID-19, and, you know, with higher exposure being linked to severity and things like that. And it just felt like [they] don’t care, they’ll just get somebody else in my shoes tomorrow.*

Staff also reported feeling underappreciated, not feeling listened to or involved in any decision-making processes, and on other occasions silenced,[c,e,h,o,q,t,v] which was often the case for members of staff from black and minority ethnic cultures.[l] Together, these findings coupled with a sense of poor decision-making by senior managers,[j,aa] a perceived culture of blame from leaders and government,[b] hierarchies perceived as worsening the working environment,[f] as well as a pressure from managers to continue working on the frontline,[l] led to participants reporting fractured relationships with managers.[g,h] A very powerful part of a longer quote from French et al.[h] reads:

> *I feel extremely frustrated, I feel powerless, I do not feel listened to, I feel like I have nowhere to go with anything. That’s why I feel like I’ve reached the end of the road. I do feel like that.*

### Theme 4: Uncertainty and Lack of Knowledge Leading to Distress and Moral Injury

Staff often reported problems related to their uncertainty and lack of knowledge in carrying out particular tasks, a phenomenon that was more intense during the first waves of the pandemic due to its novelty. They were distinct but interrelated experiences that caused distress. Staff were responsible for making challenging decisions,[a,d,p,q,v,w,x] which were perceived as distressing. A staff member reported to Liberati et al.[p]:

> *I don’t have as much information to make decisions so I’m questioning my decision-making more thoroughly. I’m frightened of making the wrong decision when I’m deciding whether somebody gets a service or doesn’t get a service. That’s quite problematic.*

This matter is unsurprising and expected from people working in health-related services, as many decisions inherently involve risk. However, there were clear systemic factors contributing to making various decisions particularly challenging. For instance, participants reported being asked to work in specialised areas about which they had limited or no training, expertise, or knowledge.[a,d,f,n,p,q,t]

The lack of established programmes, protocols or, sometimes, equipment led to anxiety and uncertainty,[d,n,q] and even participants with experience reported their lack of confidence.[s] In general, many participants were worried about harming patients due to mistakes or incorrect decisions,[d,p,q,t,v] especially when caring for people in critical conditions.[f,p,s,w,x] An illustrative quote is presented by Rees et al.[w]:

> *I feel they were pushed out too quickly without adequate training and understanding from frontline crews, and I fear this will lead to risky decisions being made that would not otherwise benefit the patients.*

Participants reported widespread concerns about the quality of care provided for patients, treating it as substandard.[f,j,m,p,q,s,v,w,x] Moreover, staff often faced dilemmas between strictly adhering to protocols and not violating their jobs’ values.[j,p,q,w] Many informants reported experiencing self-doubt, lack of control, distress, panic attacks, anxiety, guilt or moral injury due to violating their personal values and moral codes.[f,i,j,l,p,q,s,t] Grailey et al.[i] quote a staff member saying that:

> *we did absolutely the best that we could possibly do, but it just in no way, shape or form was good enough. But we did what we could in the confines of our environment.*

### Theme 5: Intense Workloads as Stressor

The last pattern that appears across the dataset in relation to stressors faced by health and social care workers is associated with workload problems. Many participants quoted in the papers available to us referred to intense and unsustainable workloads in extremely challenging and understaffed working environments.[a,b,c,d,f,i,j,n,p,q,s,t,v,z,aa] Aughterson et al.[b], for example, quote a participant stating that:

> *My routine was really like … wake up, eat something, go into work, which as shifts as nurses we had to stay in the hospital for 12 and a half hours … go home and eat something, drink something, go to sleep … then wake up and then go to work again … we have been extremely busy compared to the normality.*

Another example comes from Spiers et al.[z] who quote a participant saying:

> *… on a Friday in the middle of the day when there was no consultant around […] I gained 14 new patients who I’d not met before […] that was a really stressful day.*

Across the papers reviewed, staff from different domains of care and from various specialties were described as saying that they were overburdened by a range of additional responsibilities. They opined that their hectic environment did not allow them enough time to care appropriately for critically ill people. They experienced a sense of stagnation due to overload because of very long shifts and reported an inability to take breaks or think about how to best navigate the difficulties inherent in their jobs and especially so because of the new situation they were facing.

Associated with workload problems was the fact that staff’s roles and responsibilities were changing, as well as the requirement to adapt to redeployment and new work structures.[a,b,f,i,j,r,aa] McGlinchey et al.[r] quote a staff member who, regarding her redeployment, said:

> *That was just dropped on us. There was no negotiations, there was no ‘these are your options, you might not have to go there’ … the thought of moving again to a different hospital almost an hour away is too much for me.*

Staff clearly thought that workloads, which they perceived as excessive, also had an impact on team performance. Sometimes, workloads led to divisions and tensions between colleagues and to breakdowns in teamwork.[e,i,j,z] Being understaffed and under-resourced, for instance, led to a negative impact on team morale, which Harris et al.[j] characterise as, ‘shortage of staff; decreasing staff morale; cracks in the team.’ which, according to the same paper, was the case for the healthcare system as a whole:

> *working in hospitals that run [at] near 100% capacity near 100% of the time (prior to the outbreak) and then expecting and trying to take a service that has little slack and stretching it further. It’s been relentless and exhausting, sometimes you are left feeling that, despite doing our best, we should be doing better but can’t, given the circumstances/resources.*

The overstretched system made it hard for colleagues to care for others due to the exhaustion they were already facing. A senior doctor reported in Harris et al.[j] that:

> *My own biggest challenges have been the moral distress of watching colleagues struggle, and worrying about their wellbeing - this has been accentuated by the fact that my own world has been too busy in other related matters to be able to directly offload their workload, leading to [me] feeling inadequate for prolonged spells.*

## DISCUSSION

Our aim for the study was to provide a systematic review of qualitative papers related to the impact of the COVID-19 pandemic on UK HCWs’ experiences during the first two waves of the pandemic, which was characterised by uncertainty, high levels of infections and deaths across the globe, and the absence of vaccines. Our goal was to move beyond summarising previous results and to focus on the psychosocial effects of stressors and their particular origins. We identify five main themes from our thematic synthesis. Each theme was, mainly, centered around one pattern of stressors and a diverse range of impacts stemming from it.

The first theme concerns the direct effects of COVID-19 on healthcare workers. Due to high levels of the SARS-CoV-2 virus present in their workplaces and elsewhere, participants were worried about being infected and/or transmitting the virus to family members, friends, and acquaintances. Exposure to the virus was associated with a variable range of psychosocial outcomes including a sense of anxiety, fear, uncertainty, demoralisation, and despair. These feelings coexisted with grief and were compounded by a deliberate reduction of social contacts in order to try to reduce contamination and associated feelings of isolation. These worries were not unfounded, because higher exposure, workplace settings such as shared spaces, and being a frontline worker were associated with increased prevalence of hospitalization.[1,5]

Despite the potent direct effects of the pandemic, other stressors rooted in problematic institutional practices and arrangements were reported much more frequently in the data. For example, PPE-related problems were associated with increased rates of hospitalisation,[1] which increases worrying and over time can prove to be a significant psychological burden for healthcare workers.[4,5] Our systematic review shows that a lack of protective equipment interacted with the impact of the pandemic to intensify worry. Inability to take sufficient breaks as a result of limited access to PPE led to fatigue, dehydration, urinary infections and other health risks. Participants were worried about the quality of the PPE and the variable conduct of fit testing led to anxiety and demotivation. Many participants reported not having training and confidence in using PPE appropriately, which increased fears of exposure to the virus, and the guidance about it changed rapidly. Using PPE also caused problems in communicating with patients and with colleagues in clinical teams.

Leadership and communication were frequently reported as problematic. Rapidly changing or too much information, which was often not properly checked and was coming from too many sources led to confusion and lack of trust in both messages and sources. Senior managers were reported as largely absent from the shopfloor. Hierarchies, concerns about a perceived culture of blame, and lack of facilities led participants to feel unappreciated, undervalued, misrecognised and misunderstood, ignored, abandoned, and seen as numbers. These experiences are stressors that were present from before the pandemic [19,58] and clearly persisted, were exacerbated and exerted their negative impacts during the pandemic. Moreover, the uncertainty, which can be inherent in relatively novel events of the magnitude of the COVID-19 pandemic, was compounded by uncertainty stemming from challenging decisions and dilemmas in the absence of appropriate training, which could lead to moral distress.[2,59] Finally, excessive workloads due to chronic understaffing and staff sickness absences that predated the pandemic became unsustainable during the pandemic, and staff had to deal with continually changing roles and structures, stagnation, and, as a result, low morale, demotivation, and an impact on relationships between team members. Intense workloads and feelings of professional stagnation have also been commonly observed across large meta-analyses and have been shown to lead to negative psychosocial outcomes.[2,4,6] It appears to us that may of the themes we report were interactive and mutually reinforcing.

Apart from fear of exposure to the virus, which we regard as a primary stressor due to being inherent in the pandemic and posing an existential threat for staff and their close others, all other problems reported in our analysis either existed before the pandemic (e.g., unsustainable workloads, understaffed services, insufficient stockpiles of PPE) or were indicators and outcomes of ineffective responses to the pandemic itself (e.g., lack of training or fit testing for PPE, invisible leaders). Thus, they can be defined as secondary stressors.[10,11] We emphasise two points here. First, the effects of secondary stressors can be compounded since they can co-exist and operate in clusters, and, thereby, increase their impacts on the people affected. Second, secondary stressors can exacerbate the negative impacts of primary stressors or the pathways through which they become potent. For example, concerns regarding exposure to the virus (i.e., the primary stressor) can worsen if staff have insufficient PPE, are not trained to use it effectively, and there is uncertainty regarding its appropriate use. Uncertainty and lack of knowledge can lead to decisions that might contribute to negative outcomes for patients, increasing the sense of staff feeling personally responsible thereby exacerbating worry. Moreover, secondary stressors can amplify the effects of other secondary stressors. Being overworked can potentially lead to negative outcomes, especially in absence of established protocols due to rapidly changing guidelines. Lack of PPE erodes trust in leaders.

Overall, the notion of secondary stressors is theoretically and practically insightful as it helps us to emphasise the systemic nature of the issues raised by participants and, thus, to our ability to track these stressors and change them. In this study, secondary stressors point to occupational problems and organisational features that are problematic. In other instances (e.g., the general public), secondary stressors reflect issues related to gender and income inequality.[60] However, across all these cases, most of the negative experiences we report were not inevitable, but rather point to problems in cultures and environments in which healthcare staff work, as well as to problems arising from the wider political decisions.[10,11]

The theoretical lens of secondary stressors is also practical because it allows us to take an in-depth look into the nature of stressors and their psychosocial impacts. This framework is useful for a number of reasons. First, it helps to establish typologies of stressors as they are identified in the existing literature whose impacts can be prevented through timely identification and removal of each stressor. Second, this lens offers us transferability of insights. Although the systematic review we report in this paper records some of the lessons learned about meeting the needs of staff during the pandemic and other serious emergencies, it is clear that those lessons are also highly relevant in more ordinary times.

Thus, the usefulness of the notion of secondary stressors extends beyond the field of extreme events and into more ordinary work and workplace settings. Similar findings have come from within healthcare settings, with ambulance staff, for example, not only reporting distressing features of their work such as a lack of downtime, a target culture, and their managers and support services not being very supportive, but also identifying gaps in their training and knowledge that would improve their working conditions and professional conditions and relationships.[61]

In our experience, secondary stressors are prone to occur throughout healthcare and most other systems, and our synthesis of the experiences of HCWs during the first and second years of the COVID-19 pandemic serves as an example through which to highlight chronic problems in the NHS that promote damaging outcomes (e.g., low staff retention, problems in recruitment).

During the decade 2010-2020, the NHS struggled with staff recruitment and retention due to chronic extreme pressure caused by inequity, inequality, funding cuts, and high persisting levels of distress and fatigue. In 2023, the GMC published research that explores the reasons why doctors have left or may be considering leaving the NHS to practise abroad.[62] The report states that ‘general improvements to workplace […] could address the main reasons why doctors say they are unhappy in UK practise, and have a positive impact upon retention’. Thus, our opinion is that many of the problems with current NHS staff recruitment and retention are likely to reflect the secondary stressors faced by staff. In parallel, Maben et al. review the evidence for three workplace conditions that matter for improving quality and safety in healthcare. They regard key matters as: staffing; psychological safety, teamwork and speaking up; and staff health and wellbeing at work.[26,63] We think that these topics have much in common with the experiences we report here regarding working conditions in the NHS during the COVID-19 pandemic. Maben et al. offer helpful approaches to remedying some of the secondary stressors in healthcare services. Recent research points to the importance of social support and collective resilience in assisting frontline workers.[64] Thus, we think that what we have learned about stressors experienced by healthcare staff during the pandemic has much to teach us about improving staff retention in the NHS. Considering the dynamics and impacts of secondary stressors can help us to think more strategically about how to best support staff and care for them as well as better prepare for future extreme events.

Furthermore the concept of secondary stressors could also help us to think about limitations in other systems and institutions (e.g., schools, social care and other workplaces) including how their members cope with extreme events in the face of wider ongoing social issues (e.g., poverty, discrimination).

Lessons for Future Support

Thus, we argue that an exclusive emphasis on improving healthcare workers’ mental health through the development of personal resilience is likely to be inadequate unless it takes into account how organisational features and responses to extreme (and more ordinary) events affect healthcare workers. Our analysis of HCWs’ experiences and the stressors they faced during the early onset of the pandemic attests to that position and echoes findings from past pandemics.[65] We present a list of recommendations based on our findings that can support healthcare staff facing adverse conditions:

1. Adequate personal protective equipment (PPE) of high quality should be available.
2. Staff should be trained on how to use the equipment properly to minimise actual and perceived risks of infection and transmission.
3. Managers should plan for adequate breaks, especially for staff using PPE for prolonged hours to enable rest and reduce their fatigue.
4. Managers should streamline communications and release them from specific channels to avoid conflicting and contradictory messages. They should be communicated clearly from early on and especially so if uncertainty is expected and rapid changes occur.
5. The presence of leaders on the ground is essential in order to create a sense of team cohesion around a common purpose and so that staff feel supported and cared for. The absence of leaders can create divisions within organisations.
6. Opinions and concerns of staff should be heard and considered by managers. This is one way to increase a sense of organisational belonging and productive vertical relations.
7. Staff should be able to feel secure and supported by peers, leaders and managers when facing dilemmas and hard decisions.
8. Staff should be able to feel that they are delivering appropriate levels of care in line with the moral foundations of their profession. Otherwise, they run the risk of experiencing moral distress.
9. Healthcare institutions need appropriate levels of staffing to ensure adequate rest for staff and appropriate levels of care for patients.
10. Staff should be provided with tangible support (e.g., free parking spaces, resting facilities) which not only is likely to reduce fatigue, but will also increase team cohesion, trust in senior managers, and cultivate feelings of being cared for and supported by their institution.

## Limitations

First, we only considered papers published between 2021 and early 2022. Unavoidably, this means that significant papers of high quality that might provide novel findings that might support or contradict the findings of this review might be missing. However, even if this is the case, we consider this risk to the present analysis to be minimal because all papers from the pre-specified timeframe of the search process were in the same direction. Future research should explore healthcare workers’ experiences and stress processes in the longer-term aftermath of extreme events.

Second, the quality of each systematic review depends on the quality of the papers included in it. As expected, all the papers we reviewed have limitations. Nevertheless, we have tried to minimise the impact that these matters had on our review. We pre-registered the review with PROSPERO and followed the PRISMA and NICE guidelines on assessing the quality of the papers, the vast majority of which we judged to be of high quality. Those that did not meet this standard showed minor limitations that did not pose a risk in terms of negatively affecting our findings.

## Conclusion

A systematic review of qualitative papers published in the UK regarding the effects of COVID-19 on healthcare staff showed that, apart from primary stressors, secondary stressors were very influential in the proliferation of distress. They included inadequate leadership and communications, excessive workloads, lack of personal protective equipment, and uncertainty and lack of knowledge, each of which had a range of negative psychosocial outcomes for the people affected. Considering that recruitment and retention are continuing central concerns for the NHS, mitigation strategies should not focus so much on building individual or personal resilience in staff but rather to try to improve workplace conditions by tracking and tackling secondary stressors so that their effects are reduced.

## Data Availability

This study is a systematic review of published papers - all the studies used in the review are listed in a table in the paper itself.

## Roles of the Authors

**EN:** Led the research, worked with RW on designing the research, conducted the analysis of the papers and was instrumental in writing this paper.

**RW**: Worked with EN in designing the research, critically reviewed the themes and subthemes that emerged from the qualitative analyses and contributed to designing and writing this paper.

**KL**: Worked with EN on obtaining and analysing the papers.

**AW:** Conducted the literature search.

**AR:** Worked with EN and KL on obtaining and analysing the papers.

**All authors:** Have read and approved the final version of this paper.

