## Supplementar materials for "SECONDARY STRESSORS AND THEIR PSYCHOSOCIAL IMPACTS ON HEALTHCARE STAFF: LESSONS FROM A QUALITATIVE SYSTEMATIC REVIEW FROM THE COVID-19 PANDEMIC IN THE UK"

**Supplementary Materials**

Table 1. Mixed-methods papers not included in the systematic review

| **Authors** | **Title** | **DOI** |
| --- | --- | --- |
| **Bentham et al. (2021)** | Wellbeing of CAMHS staff and changes in working practices during the COVID‐19 pandemic | 10.1111/jcap.12311 |
| **Bhamra et al. (2021)** | Impact of the coronavirus pandemic (COVID-19) on the professional practice and personal wellbeing of community pharmacy teams in the UK | 10.1093/ijpp/riab062 |
| **Blake et al., 2021** | Psychological impacts of covid–19 on healthcare trainees and perceptions towards a digital wellbeing support package | 10.3390/ijerph182010647 |
| **Cubitt et al. (2021)** | Beyond PPE: a mixed qualitative–quantitative study capturing the wider issues affecting doctors’ well-being during the COVID-19 pandemic | 10.1136/bmjopen-2021-050223 |
| **Davies et al. (2021)** | Support for general practitioners during COVID-19 | N/A  (PMCID: PMC8581688) |
| **Dominic et al. (2021)** | ‘It’s like juggling fire daily’: Well-being, workload and burnout in the British NHS - A survey of 721 physicians | 10.3233/WOR-205337 |
| **Gemine et al. (2021)** | Factors associated with work-related burnout in NHS staff during COVID-19: a cross-sectional mixed methods study | 10.1136/bmjopen-2020-042591 |
| **Kane et al. (2021)** | The psychological effects of working in the NHS during a pandemic on final-year students: part 1 | 10.12968/bjon.2021.30.22.1303 |
| **Mitchell et al. (2021)** | Community end-of-life care during the COVID-19 pandemic: findings of a UK primary care survey | 10.3399/BJGPO.2021.0095 |
| **Yasin et al. (2021)** | The impact of the Covid-19 pandemic on the mental health and work morale of radiographers within a conventional X-ray department | 10.1016/j.radi.2021.04.008 |
